# Estimating the Incidence of Familial Exudative Vitreoretinopathy (FEVR) in The State of Michigan

**DOI:** 10.1101/2022.11.11.22282229

**Authors:** Matthew G.J. Trese, Xianggui Qu, Kimberly A. Drenser

## Abstract

**Background and Objective:** Amongst ophthalmologists, familial exudative vitreoretinopathy (FEVR) is agreed to be a rare disease. However, its incidence is currently unknown. The goal of this study is to identify the incidence of FEVR in Michigan.

**Study Design/Materials and Methods:** This retrospective chart review evaluated new patient encounters from the largest retinal specialty practice within the State of Michigan from over a 6 year period. These identified encounters were queried for the international classification of disease 10 (ICD-10) code (H35.023): exudative retinopathy, not diabetes. Review of both the individual patient encounters and the appropriate clinical imaging resulted in the identification of our FEVR cohort which was subsequently compared with the live birthrate in the State of Michigan over the same 6 years to determine an incidence rate.

**Results:** This study revealed 44 new FEVR diagnosis and 666, 102 livebirths over the 6 year period of interest. Thus, the incidence of FEVR in the state of Michigan was estimated to be 0.007%. (95% CI[0.005%- 0.009%)

**Conclusion:** Familial Exudative Vitreoretinopathy is indeed a rare diagnosis with an incidence rate of 0.007% within the State of Michigan.

## Introduction

Familial Exudative Vitreoretinopathy (FEVR) is a rare, often bilateral, inherited, vitreoretinal dystrophy originally described by Criswick and Schepens in 1969.^1^ Classically FEVR was defined by macular dragging, retinal folds, subretinal exudation and tractional retinal detachments.^2,3^ As our understanding of this disease has evolved, in large part through careful clinical observation and advancements in fluorescein angiography, we have come to understand peripheral retinal nonperfusion, retinal vascular anomalies and retinal neovascularization are driving factors for disease progression.^4-7^ Through observing these driving factors, Pendergast and Trese were able to develop a revised classification system for FEVR allowing for a unified nomenclature.^8^ This paved the way for a surgical management scheme which has helped to preserved sight for affected individuals.

More recently, several genes have been identified which when mutated are associated with the development of FEVR (NDP, FZD4, LRP5, TSPAN12, and FNZ408).^9-14^ Inheritance patterns have been reported to occur in an autosomal dominant, autosomal recessive, X-linked and sporadic manner. When juxtaposed with evidence that shows approximately 50% of patients with FEVR have no known family history, it is not surprising that phenotypic heterogeneity is common.^8,15^ In fact, mildly affected individuals will often remain asymptomatic into adulthood and may only display an avascular peripheral retina. However, severely affected individuals may progress to retinal detachment with subsequent irreversible vision loss at an early age. Due to FEVR’s high penetrance and variable expressivity, it is common to observe a spectrum of retinal findings from mild to severe within a family or even within an individual (i.e. highly asymmetric bilateral disease).

Given the high degree of variability and the wide spectrum of clinical severity, FEVR can represent a diagnostic dilemma that is not always easily recognized. While most ophthalmologists will agree that FEVR is a rare disease, its incidence remains unknown. This issue is compounded by the fact that FEVR does not have its own diagnosis code which precludes analysis by large data sets, such as the Intelligent Research in Sight (IRIS) Registry. This study aims to take a fundamental step towards understanding this patient population by estimating the incidence of FEVR within the State of Michigan over a 6 year period.

## Patients/Materials and Methods

To define our study cohort (see Figure 1), a review of the electronic health record for all physicians at Associated Retinal Consultants (ARC) was performed from January 1^st^, 2015 – December 31^st^, 2020. This identified all patient encounters for the period of interest. Next, we refined our dataset by excluding established patients (or previously examined patients), patients who did not have their primary domicile within the State of Michigan and patients whose referring doctor was from outside the United States. Next, the identified patient records were queried for the International Classification of Disease 10 (ICD-10) code exudative vitreoretinopathy, not diabetes (H35.023). Because H35.023 is not a specific ICD-10 code for FEVR, individual patient records were analyzed to ensure our cohort consisted exclusively of patients diagnosed with FEVR. Therefore, all individuals whose diagnosis was an exudative vitreoretinopathy other than FEVR were excluded from analysis. Finally, the remaining patient records were cross referenced with our widefield imaging database which used multiple imaging systems (Optos 200TX & Optos California, Optos inc., Dunfermline, UK, Poenix ICON, Phoenix Technology Group, LLC, Pleasanton, CA, Retcam2, Clarity Medical Systems) over the 6-year period of interest. We excluded any patients whose WFA was not consistent with the diagnosis of FEVR. Finally, we drew upon publicly available birth data that is compiled and reported by the State of Michigan on an annual basis to assist with our calculation of the incidence of FEVR.^16^

**Figure 1:**
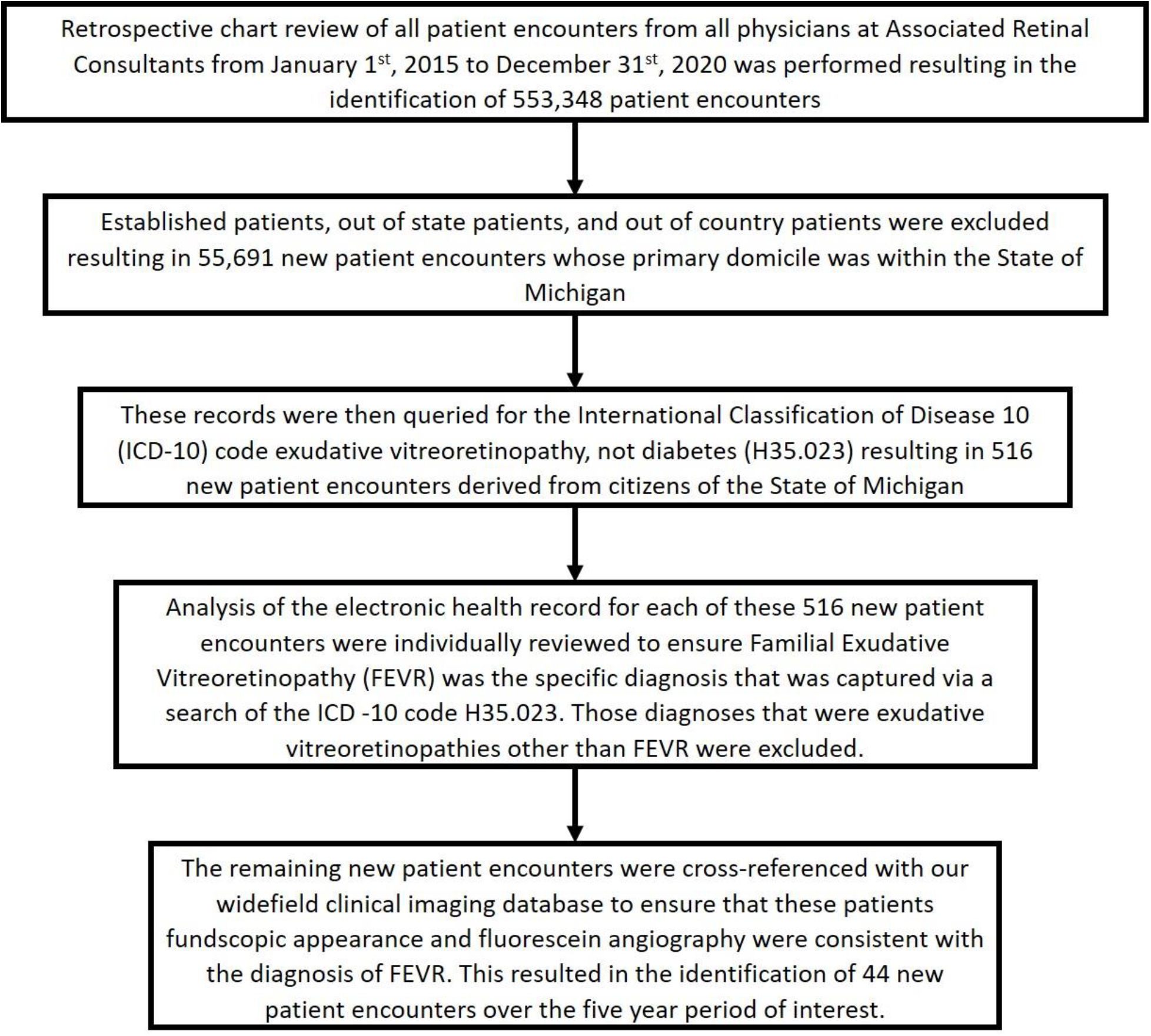
Identifying Familial Exudative Vitreoretinopathy Patient Encounters. The figure above outlines the steps taken to identify the charts of individuals residing in Michigan that were newly diagnosed with FEVR from 2015-2020 at Associated Retinal Consultants.

These data were analyzed and an estimation of the incidence of FEVR within the State of Michigan was performed.

## Results

Over the six-year period of interest, there were 553,348 patients examined at ARC at across the state of Michigan. Of these, 55,691 were new patient visits. Within this subgroup, 516 patient records were identified which were associated with the ICD-10 code exudative vitreoretinopathy, not diabetes (H35.023). After review of the charts, 336 patients were identified as having FEVR and 134 were Michigan residents. The majority were established patients, with 44 new patients that both carried the H35.023 code, were diagnosed as FEVR, confirmed by characteristic FA findings. The ages of these newly diagnosed FEVR patients who resided in Michigan ranged from less than 1 year old to 67 years old; with the average age of FEVR diagnosis being 10 years old. Of these newly diagnosed FEVR patients, 15.9% (7/44) of these individuals were diagnosed as a direct result of a screening exam that occurred after a family member was diagnosed with FEVR. (Table 1).

The majority of new patients with FEVR (32/44) were referred to 3 physicians in the practice who specialize in pediatric vitreoretinal surgery. The remaining 12 patients were referred to the other physicians in the practice. Nine patients were seen in the southeast Michigan region (10 physicians; 37,419 new patients), 1 in western Michigan (3 physicians; 5,826 new patients), and 2 in northern Michigan (4 physicians; 12,481 new patients). This equates to 0.11% in southeast Michigan and 0.02% in both western and northern Michigan. When the three pediatric vitreoretinal surgeons were removed the southeast Michigan office had a similar rate to the other regions of 0.03%, suggesting that the majority of patients with retinal exams suggestive of FEVR are referred to a specialist in that field. In fact, the 3 specialist saw 7,984 new patients during that 6 year window and 41 were diagnosed with FEVR, equating to 0.5%.

Next, we accessed publicly available birth data published by the State of Michigan. The birthrate, has remained relatively stable over the last 50 years, including the six-year period of interest and the 10 prior years. Over the six-year period of interest there were 666,102 live births. Thus the estimated incidence of FEVR in Michigan over the six-year period was 0.007% (44/666,102). If we assume that the 44 FEVR patients represent a random sample of FEVR patients from within 666,102 individuals, then we are 95% confident that the incidence of FEVR is 0.005%-0.009%.

## Discussion

As expected, our study confirms that FEVR is a rare disease as designated by the U. S. Food and Drud Agency (prevalence less than 200, 000 in the United States). Most retinal specialists will see 0-1 patient with FEVR over six years, or roughly 1 in 5,000 patients (0.02%), based on the data from the 17 retinal specialists in our practice. Through evaluation of our new patients’ charts, as well as the relevant imaging (including fundus photography and WFA) over a six-year period, we determined the incidence of FEVR within the State of Michigan to be 0.007%. We recognize that these data reflect the incidence of FEVR within a single retinal specialty practice. However, ARC physicians account for approximately 1/3 (20 out of 61) of the retinal physicians in the State of Michigan. Further, ARC specializes in pediatric retinal disease and is a well-recognized national and international tertiary referral center for FEVR as well as other rare pediatric retinal diseases. Thus, we expect that our analysis captures the vast majority of the pediatric retinal disease referrals within the State. We recognize that our incidence estimate may not be entirely comprehensive because we are reporting data solely from our own experience. Despite this, we recognize that FEVR, particularly in its early stages, can represent a diagnostic challenge. This particularly true for ophthalmologists who are not attune to the spectrum of clinical findings associated with FEVR and who are not implementing the regular use of WFA for suspected patients. Because of our experience with the evaluation, diagnosis and management of this rare disease we feel that our incidence estimate may represent a modest overestimation of the true incidence of FEVR within Michigan.

To our knowledge, only one other study has indirectly estimated the incidence rate of FEVR.^17^ In this multicenter, cross-sectional study, the authors evaluated nearly 200,000 Chinese newborns to assess the utility of universal fundus screening using fundus photography. They reported that nearly 10% of newborns screened demonstrated an abnormal fundus finding, most commonly a retinal hemorrhage. However, the authors also reported that over the 8-year period there were 217 cases of FEVR which accounted for 1.19% of all abnormalities and greatly outnumbering the prevalence of Retinopathy of Prematurity (ROP), a finding that is inconsistent with our clinical experience. Over the same six-year time period, 552 premature infants were examined for ROP in the southeast Michigan office alone. The Early Treatment-ROP study found 39% of premature infants to have retinopathy, suggesting 215 infants with clinically significant ROP were seen in the southeast Michigan hub compared to 41 FEVR patients over the same six-year time period. While we applaud the authors for their use of universal screening with fundus photography, this estimation of the incidence of FEVR is several orders of magnitude higher than our estimation. This is likely accounted for by the methodology used to identify the patients and the lack of diagnosis confirmation by WFA. FEVR can not be properly diagnosed without the use of WFA Previous research has shown that fluorescein angiography greatly aids in arriving at the diagnosis of FEVR through highlighting fundus findings which may be imperceptible on clinical examination (or fundus photography) alone.^7^ We feel that by taking the time to perform a comprehensive chart review, which included both review of the electronic health record as well as a thorough analysis of widefield imaging, we significantly strengthened our diagnostic precision and thus improved the accuracy of our incidence calculation.

By estimating the incidence of FEVR within the State of Michigan we have gained valuable insight into this potentially vulnerable population. Rare diseases, by the nature of being rare, are often under studied and poorly followed. This data provides a starting point for uncovering the incidence and prevalence of a rare disease that can have devastating effects on a persons productivity and quality of life. Furthermore, early intervention can result in preserving meaningful vision but requires ophthalmologists to be aware of the condition and its frequency within the general population. Further research is required to gain a more complete estimate of the incidence and prevalence of FEVR throughout the United States and the World.

## Data Availability

All data produced in the present work are contained in the manuscript

## Notes

**Grant Support, Disclosures and Previous Presentation:** The authors have no received grant funding for this project. They report no relevant financial disclosures. The content of this work has not been previously submitted or presented elsewhere.

### Competing Interest Statement

The authors have declared no competing interest.

### Funding Statement

None

### Author Declarations

WCG IRB of Associated Retinal Consultants gave ethical approval for this work

